# What Urine Measures Is Not What Tissue Encodes: Compartment-Specific miRNA Coordination in Prostate Cancer

**DOI:** 10.64898/2026.06.14.26355623

**Authors:** Shweta Singh, Pramit Biswas, Sameer Trivedi, Mahima Yadav, Manjari Gupta, Lalit Kumar, Yashasvi Singh, Ujwal Kumar, Parimal Das, Garima Jain

**Author notes:** These authors contributed equally to this work (Shweta Singh, Pramit Biswas).

## Abstract

1

**Background:** Prostate cancer (PCa) diagnosis remains challenged by the limited specificity of prostate-specific antigen (PSA) testing, which cannot reliably distinguish malignancy from benign prostatic hyperplasia (BPH). MicroRNAs (miRNAs) are emerging candidates for liquid biopsy-based diagnostics, but most studies assess expression in isolation within a single compartment (biological source - Tissue, blood, serum, urine etc.), overlooking both compartment-specific behavior and the coordinated relationships among miRNAs.

**Methods:** We profiled four candidate miRNAs — miR-19b-3p, miR-21-5p, miR-101-3p and miR-375-3p, across four biological compartments (prostate tumor tissue, urine, serum, and blood) in 179 patients undergoing prostate biopsy for clinical suspicion of PCa (104 PCa, 75 BPH) using qRT-PCR. Urinary exosomal RNA was isolated with a commercial exosome isolation kit so from here onwards this compartment will be referred to as urine. Differential expression was quantified using Cohen’s *d*; inter-miRNA coordination was assessed via Spearman correlation and differential correlation (Δ*r*) analysis; and a compartment-level network rewiring score was derived as the sum of |Δ*r*| across miRNA pairs. Cross-compartment structural alignment was evaluated by comparing correlation patterns at the population level. Diagnostic models combining PSA, age, and urinary exosomal-miRNA features were evaluated using Logistic Regression, Elastic Net Logistic Regression and Naive Bayes classifiers under leave-one-out cross-validation (LOOCV).

**Results:** Effect sizes were largest and most consistent in urine, with miR-101-3p showing the strongest separation between PCa and BPH (*d* = −1.01), followed by miR-21-5p (*d* ≈ −0.72) and miR-19b-3p (*d* ≈ −0.64). Two markers (miR-19b-3p, miR-375-3p) showed directional reversals across compartments, indicating that disease-associated signals are compartment-specific rather than uniformly conserved. In tumor tissue, PCa was associated with substantial reorganization of inter-miRNA coordination (network rewiring score = 2.46), including the emergence of a strong miR-21-5p–miR-375-3p co-regulatory axis (Δ*r* = +0.87) and decoupling of the miR-21-5p–miR-19b-3p relationship (Δ*r* = −0.64). Urine showed a structurally distinct coordination pattern (rewiring score = 1.77), dominated by a miR-101-3p–miR-19b-3p axis (Δ*r* = +0.56) absent from tissue; cross-compartment comparison showed concordance in only 1 of 6 miRNA pairs, indicating that urine’s architecture is largely independent of tissue’s.

For diagnostic translation, the conventional PSA cutoff (4 ng/mL) achieved 100% sensitivity but only 23.5% specificity. In urine, miR-101-3p performs better than other miRNAs, with AUC of 0.71 (95% CI: 0.56–0.85). Adding PSA and age to the urinary miR-101-3p further improved discrimination to an AUC of 0.91 (95% CI: 0.82–0.99), with 70.5% specificity at 92.8% sensitivity; this pattern was consistent across Elastic Net and Logistic Regression classifiers. Expanding the model to include all urinary miRNAs, age, and pair-derived coordination features did not improve on this result (AUC = 0.88), indicating that population-level coordination changes did not translate into additional individual-level diagnostic value in this cohort.

**Conclusions:** miRNA signals in extracellular compartments do not represent direct surrogates of tumor-level molecular architecture; each compartment harbors a distinct, transformed coordination structure reflecting its biological context. While these coordination-level changes are mechanistically informative, the most direct translational gain in this study came from a parsimonious model combining PSA, age with a single urinary marker, miR-101-3p, which improved AUC from 0.71 to 0.91, with specificity 70.5% at 90% sensitivity criteria. This combination represents a promising, interpretable candidate for reducing unnecessary prostate biopsies, pending validation in larger, independent cohorts.

## 2 Introduction

Prostate cancer (PCa) remains one of the most commonly diagnosed malignancies in men and a leading contributor to cancer-related mortality worldwide [1, 2]. Early detection is central to improving outcomes, yet the diagnostic tools currently available carry significant limitations [3]. Prostate-specific antigen (PSA) testing remains the standard screening approach, but its lack of specificity is well documented: PSA levels rise not only in malignancy but also in benign prostatic hyperplasia (BPH), prostatitis, and other non-cancerous conditions common in aging men [4]. Furthermore, approximately 40 % of mp-MRIs are also reported as uncertain [5]. Thus the current diagnostic process leads to a substantial proportion of unnecessary biopsies, with associated procedural risk, patient anxiety, and healthcare cost. Closing this diagnostic gap requires biomarkers that are minimally invasive, reproducible, and capable of distinguishing malignant from benign prostate pathology with greater confidence than PSA alone provides.

MicroRNAs (miRNAs) have emerged as compelling candidates to fill this gap. These short, non-coding RNAs regulate gene expression post-transcriptionally and are remarkably stable in biological fluid resistant to the enzymatic degradation that limits many other RNA-based markers [6]. They are detectable in tumor tissue as well as in circulating biofluids such as serum, whole blood, and urine, making them accessible through routine, minimally invasive sampling [6]. A growing body of literature has linked specific miRNAs including miR-21-5p, miR-19b-3p, miR-375-3p, and miR-101-3p to prostate cancer diagnosis and progression across multiple sample types [7–11].

Despite this promise, no miRNA-based assay has reached routine clinical use for PCa diagnosis, and results across studies remain notably inconsistent. While this variability is widely attributed to technical and clinical heterogeneity, it may be further compounded by an unresolved biological question that has received surprisingly little systematic attention: compartment specificity (word compartment represent biological source — tissue, blood, serum, urine etc.). Most biomarker studies examine a single biofluid in isolation, say, serum or urine. A second, related blind spot concerns how biomarker studies typically define “signal”. Conventional differential expression analysis asks a single question: is a given miRNA present at higher or lower levels in disease versus control? This captures real and useful information, but it is only one dimension of what biological systems encode. Genes and their regulatory products do not act in isolation they form coordinated networks, and disease processes often reorganize the relationships between molecules even when the abundance of any single molecule changes only modestly [12, 13]. This phenomenon, broadly described as network rewiring, has been documented across cancer biology: a strong correlation between two molecules in normal tissue may weaken, disappear, or reverse in malignancy, signaling a shift in shared regulatory control [14–16]. For miRNAs specifically, this means that two markers could show nearly identical average expression in PCa and BPH while differing substantially in how tightly they co-vary a pattern invisible to standard expression-based analyses [15, 17].

These two issues compartment specificity and molecular coordination are not independent. If disease-associated expression changes differ across compartments, it stands to reason that coordination patterns might differ too. A co-regulatory relationship observed in tumor tissue may not survive the journey into the bloodstream or urine; conversely, extracellular compartments may develop their own coordination architectures, shaped by the biology of secretion, vesicle packaging, and clearance, that are distinct from, but not necessarily less informative, than what is seen in the tumor itself.

This raises a chain of questions that motivate the present study. Are PCa-associated miRNA changes consistent across tissue, blood, serum, and urine, or are they compartment-specific. Does cancer-driven network rewiring observed in tumor tissue propagate into extracellular compartments, or do liquid biopsy signals represent something more transformed? And specifically for urine the most clinically convenient of the non-invasive compartments does it retain biologically meaningful information about tumor organization despite the processes of vesicle packaging, release, filtration, and excretion that intervene between tumor and sample?

Answering these questions matters beyond basic biology. If miRNA coordination encodes disease-relevant information that expression levels alone do not capture, then network-derived features could provide diagnostic value alongside conventional expression panels. Machine learning approaches are well suited to testing this possibility, as they can integrate expression-based and coordination-based features simultaneously and evaluate their combined contribution to disease classification.

To address these questions, this study profiled the four candidate miRNAs across tumor tissue, whole blood, serum, and urine in patients undergoing biopsy for suspected PCa, classified as PCa or BPH on histopathology. By combining differential expression analysis, differential correlation analysis, and cross-compartment comparison, with urine receiving particular emphasis as the most clinically accessible compartment.

## 3 Materials and methods

### 3.1 Study Population

This study was conducted at MIRNOW Lab, BioNest-Banaras Hindu University (BHU), in collaboration with the Department of Urology and Department of Pathology at the Institute of Medical Sciences (IMS) BHU, Department of Computer Science, BHU, and Centre for Genetic Disorder, BHU. From November 2022 to March 2025, we collected 179 biological samples, which included 45 urine samples, 60 whole blood samples, 42 serum samples, and 32 prostate tissue samples from patients attending the Department of Urology, IMS-BHU. The dataset included 42 serum samples (22 PCa, 20 BPH), 45 urine samples (28 PCa, 17 BPH), 32 tissue samples (20 PCa, 12 BPH), and 60 blood samples (34 PCa, 26 BPH). Samples were independent across compartments, with no overlap between them. The clinical characteristics details of patients across different biological samples stratified by disease status has given in Table 1.

**Table 1:** Clinicopathological Features of Patients in the Multi-Compartment PCA and BPH Cohorts.

| Sample Type | Group | No. of Samples | Age (Mean ± SD) | PSA (ng/mL), Range |
| --- | --- | --- | --- | --- |
| Overall | All Subjects | 179 | 65.66 ± 11.70 | 5–100 |
| | PCA | 104 | 67.45 ± 11.40 | 11–100 |
| | BPH | 75 | 63.30 ± 11.77 | 6–74 |
| Blood | All Subjects | 60 | 66.95 ± 10.55 | 13–100 |
| | PCA | 34 | 70.43 ± 10.10 | 13–100 |
| | BPH | 26 | 63.30 ± 9.97 | 17–53 |
| Serum | All Subjects | 42 | 61.62 ± 12.69 | 9–87 |
| | PCA | 22 | 64.68 ± 12.98 | 21–87 |
| | BPH | 20 | 58.25 ± 11.77 | 9–48 |
| Tissue | All Subjects | 32 | 68.28 ± 10.59 | 6–100 |
| | PCA | 20 | 67.90 ± 10.28 | 23–100 |
| | BPH | 12 | 68.92 ± 11.51 | 6–59 |
| Urine | All Subjects | 45 | 66.40 ± 11.92 | 4–68 |
| | PCA | 28 | 67.07 ± 11.77 | 17–68 |
| | BPH | 17 | 65.29 ± 12.44 | 11–46 |

Patients scheduled for prostate biopsy due to clinical suspicion of prostate cancer based on elevated PSA levels and/or abnormal digital rectal examination (DRE) findings and/or mpMRI were enrolled in the study (Tab. 1). The Ethics Committee of the Institute of Science, BHU reviewed and approved the study protocol (Registration No.: ECR/226/Indt/UP/2014/RR22; Approval No.: I.Sc./ECM-XIV/2022-23). The written informed consent was taken from all participants prior sample collections. Patients having a previous history of inflammatory diseases, kidney stones, or any malignancy were excluded from the study. Based on histopathological examination of 12-core prostate biopsy specimens, participants were categorized into two groups: BPH and PCa.

### 3.2 Sample Collection and Storage

About 15 mL of urine was collected in sterile containers. 1 ml whole blood samples were collected in EDTA coated tubes and 2 ml in clot activator tubes for serum. Prostate tissue samples were collected in RNAlater (Thermo Fisher Scientific, USA). Blood samples were aliquoted into microcentrifuge tubes to be stored at −80°C until further processing. For serum preparation, blood samples in clot activator tubes were centrifuged at 10,000 rpm for 10 minutes at 4°C. We collected the supernatant and stored it at −80°C until needed. Prostate tissue and urine samples were also stored at −80°C.

### 3.3 RNA Isolation from Blood, Tissue, Serum, and Urine

Total RNA from whole blood, serum and tissue using the TRIzol-chloroform extraction method. The RNA isolation protocol for blood [11], tissue [18], and serum [19] followed previously detailed methods. We used the Norgen Urine Exosome RNA Isolation Kit (Cat. No. 47200, Norgen Biotek Corporation, Canada) to isolate urinary exosomal RNA according to the manufacturer’s instructions 10 mL of clarified urine following the recommended protocol manual. RNA concentration and purity were assessed using a NanoDrop OneC UV–Vis Spectrophotometer (Thermo Fisher Scientific, USA).

### 3.4 cDNA Synthesis and Quantitative Real-Time PCR (qRT-PCR)

cDNA synthesis using the First Strand cDNA Synthesis Kit (Thermo Scientific, Catalog No. K1622, USA). Both total RNA and exosomal RNA were reverse-transcribed with miRNA-specific stem-loop primers. Details of the primer sequences are in Supplementary Table 1. Before qRT-PCR analysis, cDNA diluted at a ratio of 1:5 with nuclease-free water. qRT-PCR experiment was performed using the QuantStudio™ 6 Flex Real-Time PCR System (Applied Biosystems, USA). Each 10 *µ*L PCR reaction included 1 *µ*L of diluted cDNA, 1 *µ*L each of forward and reverse primers (10 *µ*M), 5 *µ*L Maxima SYBR Green/ROX qRT-PCR Master Mix (2X Thermo Scientific, Catalog No. K0221), and nuclease-free water to makeup the final volume. The thermal cycling conditions started with an initial denaturation step at 95°C for 10 minutes, followed by 40 amplification cycles of 95°C for 15 seconds and 60°C for 1 minute. The relative gene expression analysis using the comparative Ct value.

### 3.5 Data Description

The dataset consisted of patient samples categorized into two clinical groups: PCa and BPH. miRNA expression levels were measured as cycle threshold (Ct) values using qRT-PCR across four biological compartments: serum, urine, tissue, and blood. The analyzed miRNAs included miR-21-5p, miR-19b-3p, and miR-375-3p across all compartments, with an additional miR-101-3p measured in urine and tissue samples. RNU6 was used as normalizer in blood, tissue and serum, whereas let-7c was used as a normalizer for urine exosome samples which corresponds to the previously published data[20]. dCt values represent inverse measures of gene expression, where lower dCt values indicate higher expression levels.

### 3.6 Data Preprocessing

Data was preprocessed to ensure consistency and reproducibility, column names were standardized across all sheets. Clinical group information was stored in a column labeled groups, indicating the diagnostic category of each sample. For machine learning applications, categorical labels were encoded numerically, with BPH assigned a label of ‘0’ and PCa assigned a label of ‘1’. Missing values were handled using a group-wise median imputation strategy. Imputation was applied only to numerical variables, ensuring that the statistical properties of each clinical group were preserved while minimizing bias introduced by global imputation. Since Ct values are inversely proportional to gene expression, all Ct values were transformed by multiplying them by −1 prior to statistical analysis. This transformation ensured that higher values correspond to higher expression levels, thereby enabling more intuitive interpretation of expression differences and correlation patterns.

### 3.7 Statistical Analysis

Descriptive statistics were computed separately for each miRNA within each biological compartment (serum, urine, tissue, and blood) and clinical group (PCa and BPH). To quantify the magnitude of expression differences, Cohen’s *d* effect size was computed for each miRNA within each compartment (Suppl. Method). Effect sizes were computed only when sufficient observations were available in both groups. To investigate changes in gene–gene relationships, Spearman rank correlation coefficients were computed for each gene pair separately within PCa and BPH groups, denoted as *r*_PCa_ and *r*_BPH_. The difference in correlation was calculated as:

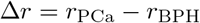

(Suppl. Method).

Additionally, variance of each gene within PCa and BPH groups was computed to evaluate expression stability across conditions. Adjusted p-values (or q-values) below the predefined significance threshold were considered statistically significant(Not reported here).

The extent of disease-associated changes in miRNA coordination was quantified using two complementary network metrics derived from pairwise correlation differences (Δ*r*) between the PCa and BPH groups. First, a Network Rewiring Score (NRS) (Suppl. methods) was calculated as the sum of the absolute correlation changes across all miRNA pairs. Second, a Cross-Compartment Concordance Score (CCCS) was used to compare rewiring patterns between tissue and urine. CCCS was computed as the cosine similarity (Suppl. methods) between the vectors of (Δ*r*) values from the two compartments (Suppl. methods).

### 3.8 Feature Engineering

To capture relative expression dynamics, pairwise and ratiowise features were constructed based on intra-compartment gene relationships. For each gene pair (*g*_1_, *g*_2_), a feature was defined as

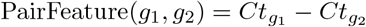

These features represent relative expression differences and can provide more robust discriminatory information than individual gene measurements. In addition to expression-based features, correlation-based features were derived to capture interaction changes between clinical conditions. For each gene pair, the absolute correlation difference |Δ*r*| was used as a measure of interaction rewiring.

### 3.9 Machine Learning Models

To classify samples into PCa and BPH groups supervised machine learning models were evaluated including Logistic Regression (LR), Elastic Net Logistic Regression (EN) and Naive Bayes (NB). These models represent a range of learning paradigms, including linear, probabilistic, distance-based, and ensemble approaches, enabling comprehensive evaluation of classification performance.

### 3.10 Evaluation Metrics

Model performance was assessed using standard classification metrics, including accuracy, sensitivity, specificity, and the area under the receiver operating characteristic curve (ROC–AUC). These metrics provide complementary insights into classification performance, particularly in biomedical contexts. Leave One Out Cross Validation (LOOCV) was used for maximizing the training data per fold (*n* −1 in *n* samples) for threshold tuning and unbiased performance estimation in small cohorts.

### 3.11 System Requirement

All analyses were implemented in Python. Data preprocessing and manipulation were performed using pandas and NumPy, statistical analysis using SciPy, and machine learning using scikit-learn. Visualization was conducted using Matplotlib and Plotly. Experiments were carried out in a Python development environment such as Jupyter Notebook or Visual Studio Code.

## 4 Results

### 4.1 Compartment-Specific Expression Patterns

To assess whether miRNA expression changes associated with prostate cancer are consistent across biological compartments we analysed Cohen’s D effect size (d-value). Effect size quantifies the magnitude of differentiation capacity of a biomarker between PCa and BPH within a specific biological compartment. This analysis was performed across tissue, serum, blood, and urine-exosomes (will be mentioned as urine only in next sections) for each of the four candidate miRNAs. The data was collected as quantification of the expression levels of selected miRNAs using qRT-PCR.

Substantial heterogeneity in effect size was observed across both markers and compartments (Fig. 1). Negative *d*-value indicates lower expression in PCa relative to BPH. The largest and most consistent effects detected in urine for miR-101-3p (*d* = −1.01), and miR-21-5p (*d* ≈ −0.72) representing a large effect by conventional standards. Additionally, miR-19b-3p (*d* ≈ −0.64) from the urine compartment also showed moderate-to-large effects, reinforcing urine as the most discriminative compartment overall (Fig. 1).

**Figure 1:**
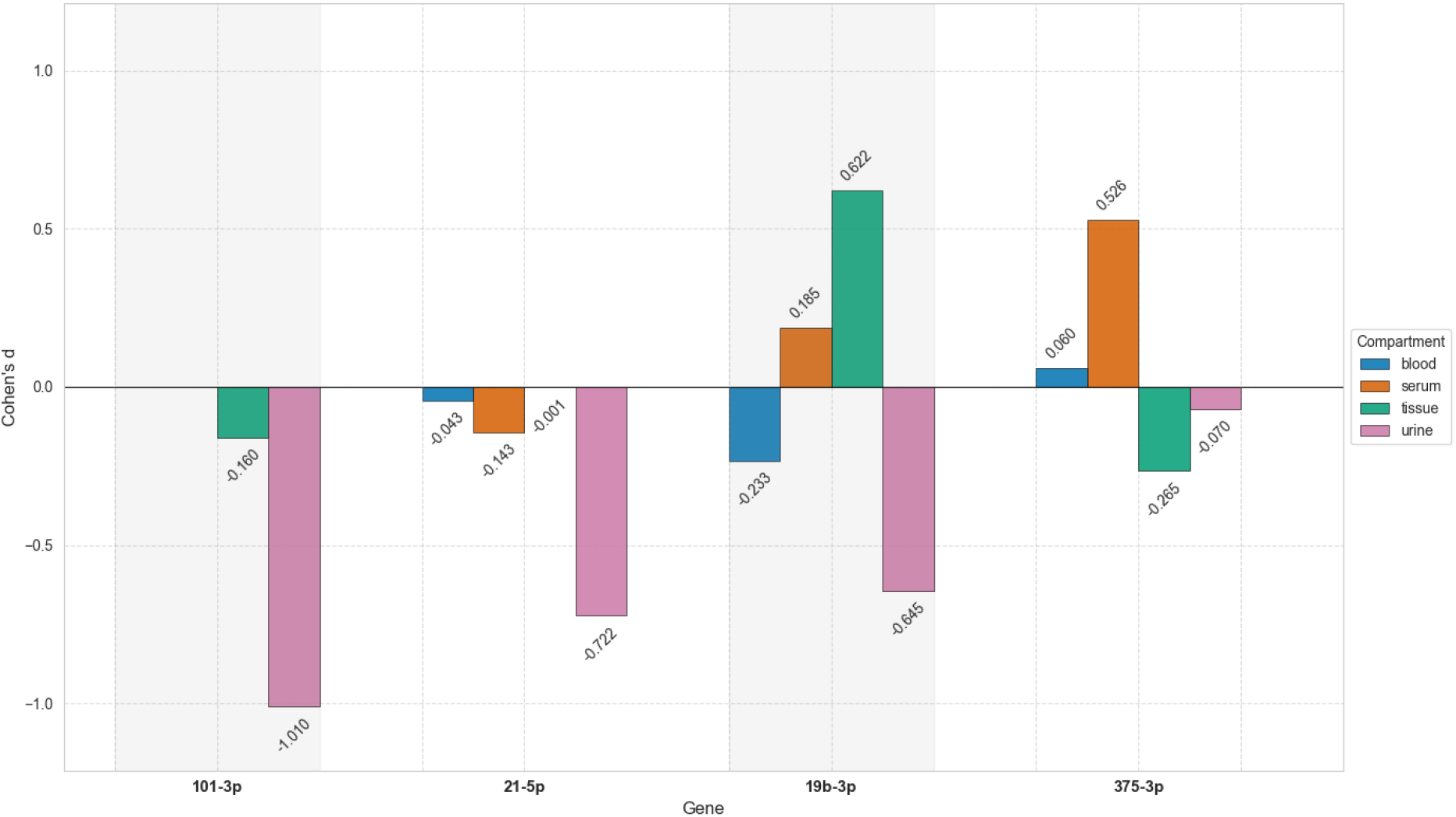
Compartment-specific effect sizes of candidate miRNAs across tissue, serum, blood, and urine. Negative values indicate lower expression in PCa relative to BPH, whereas positive values indicate higher expression in PCa.

Tissue and serum showed more selective, marker-specific patterns rather than a uniform signal. miR-19b-3p demonstrated a moderate positive effect in tissue (*d* ≈ 0.62), while miR-375-3p also showed a moderate positive effect in serum (*d* ≈ 0.53). Blood, in contrast, showed consistently small effect sizes across all four markers (|*d*| *<* 0.25), indicating minimal separation between PCa and BPH regardless of which miRNA was examined (Fig. 1).

Critically, effect sizes were not merely inconsistent in magnitude; several markers showed outright reversals in direction across compartments. miR-19b-3p exhibited positive effects in tissue and serum but negative effects in blood and urine. miR-375-3p followed a similar pattern, with negative effects in tissue but positive effects in serum.

Taken together, these results indicate that disease-associated miRNA signals are not uniformly expressed across biological compartments. Instead, both the magnitude and direction of PCa-associated expression changes are compartment-specific, with urine capturing the most pronounced and consistent discriminative signal among the four compartments examined.

### 4.2 Diagnostic Performance Mirrors Compartment-Specific Expression

Having established that effect sizes differ markedly by compartment, we next assessed whether this translated into differences in univariate diagnostic performance. ROC analysis was performed for each miRNA within each compartment, with PCa versus BPH classification as the outcome. In urine, miR-101-3p achieved the highest discriminative performance of any marker-compartment combination (AUC ≈ 0.77). miR-21-5p and miR-19b-3p in urine also showed acceptable discrimination (AUC ≈ 0.69 and ≈ 0.68, respectively), while miR-375-3p performed near chance in urine (AUC ≈ 0.54). All the patterns of AUC values were consistent with effect size findings (Fig. 2).

**Figure 2:**
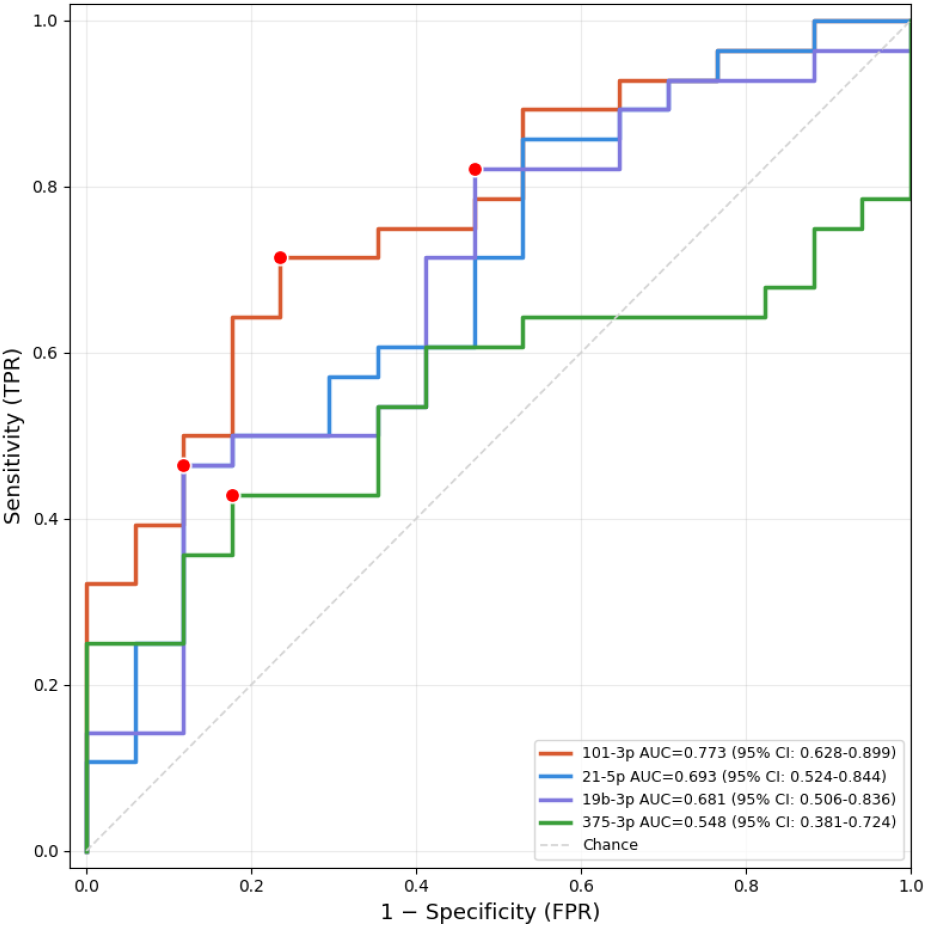
Diagnostic performance of urinary miRNAs for distinguishing prostate cancer from benign prostatic hyperplasia. Receiver operating characteristic (ROC) curves showing the classification performance of individual urinary miRNAs. Area under the curve (AUC) values and corresponding 95% confidence intervals are shown in the legend.

Serum, blood and tissue showed inconsistent performance across markers, without a single marker reaching the discriminative level (Suppl. Fig.1 (a),(b),(c)). miR-19b-3p reached AUC ≈ 0.30 in tissue and AUC ≈0.50 in serum, while miR-375-3p reached AUC ≈ 0.46 in serum but no marker performed well across all compartments simultaneously.

Overall, only urine provided consistent discrimination across multiple markers, while serum and tissue showed marker-dependent and inconsistent performance, and blood showed negligible diagnostic value regardless of marker.

### 4.3 Prostate Cancer Reorganizes Inter-miRNA Coordination

The expression and diagnostic analyses presented above characterize each miRNA as an independent feature. However, biological systems function through coordinated regulatory relationships rather than isolated components, and disease can reorganize these relationships even when individual expression levels change only modestly. Tumor tissue is the primary source of cancer-associated molecular signals, we therefore asked whether the relationships between the four candidate miRNAs differ between malignant and benign cases. To address this, we performed differential correlation analysis, comparing pairwise Spearman correlation coefficients between PCa and BPH groups for each of the six possible miRNA pairs (Tab. 2).

**Table 2:**
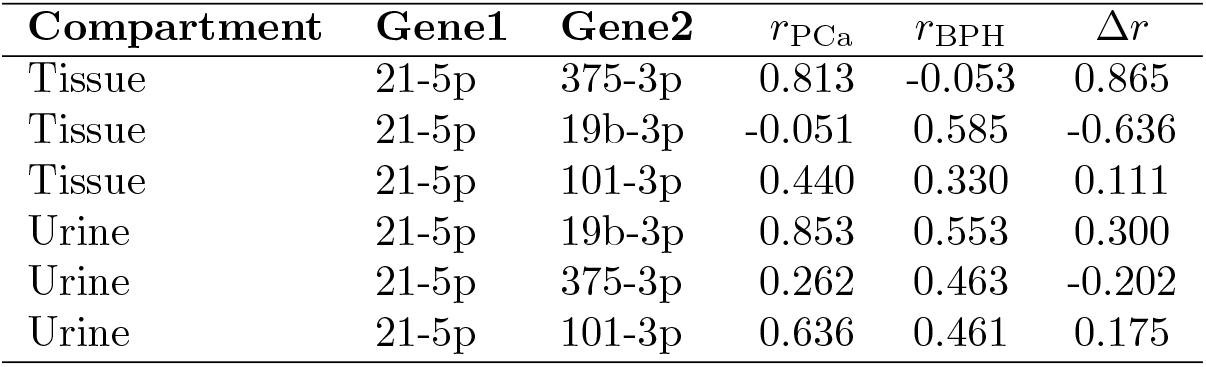
Differential correlation analysis of selected miRNA pairs in tissue and urine compartments. Pairwise Spearman correlation coefficients were calculated separately for PCa and BPH groups. Differential correlation (Δr) was defined as r_PCa_ − r_BPH_. Positive Δr values indicate strengthening of coordination in PCa, whereas negative values indicate weakening or loss of coordination.

| Compartment | Gene1 | Gene2 | $r_{PCa}$ | $r_{BPH}$ | $\Delta r$ |
| --- | --- | --- | --- | --- | --- |
| Tissue | 21-5p | 375-3p | 0.813 | -0.053 | 0.865 |
| Tissue | 21-5p | 19b-3p | -0.051 | 0.585 | -0.636 |
| Tissue | 21-5p | 101-3p | 0.440 | 0.330 | 0.111 |
| Urine | 21-5p | 19b-3p | 0.853 | 0.553 | 0.300 |
| Urine | 21-5p | 375-3p | 0.262 | 0.463 | -0.202 |
| Urine | 21-5p | 101-3p | 0.636 | 0.461 | 0.175 |

Several miRNA pairs showed substantial changes in correlation structure between the two conditions. The most pronounced shift was observed between miR-21-5p and miR-375-3p in the tissue compartment. In BPH tissue, these two markers showed *r* = −0.05, indicating that they vary independently of one another in benign tissue. In PCa tissue, however, the same pair showed a strong positive correlation (*r* = +0.81), achieving a differential correlation of Δ*r* = +0.87 (Tab. 2, Fig.3). A second notable change involved miR-19b-3p and miR-101-3p in urine. In BPH, this pair showed a moderate positive correlation (*r* = +0.14). In PCa, the correlation increased to *r* = 0.70, suggesting these two oncomiRs strongly co-vary under malignant conditions; corresponding to a large positive differential correlation (Δ*r* = 0.56) (Tab. 2, Fig.3). Interestingly, the miR-101-3p and miR-375-3p pair in urine showed slight reversal in relationship. This pair showed a modest positive correlation (Δ*r* ≈ 0.25) in BPH versus a slight anti-correlation in PCa (*r* ≈ −0.14) (Tab. 2, Fig.3). The remaining pairs showed smaller shifts (data not shown here).

**Figure 3:**
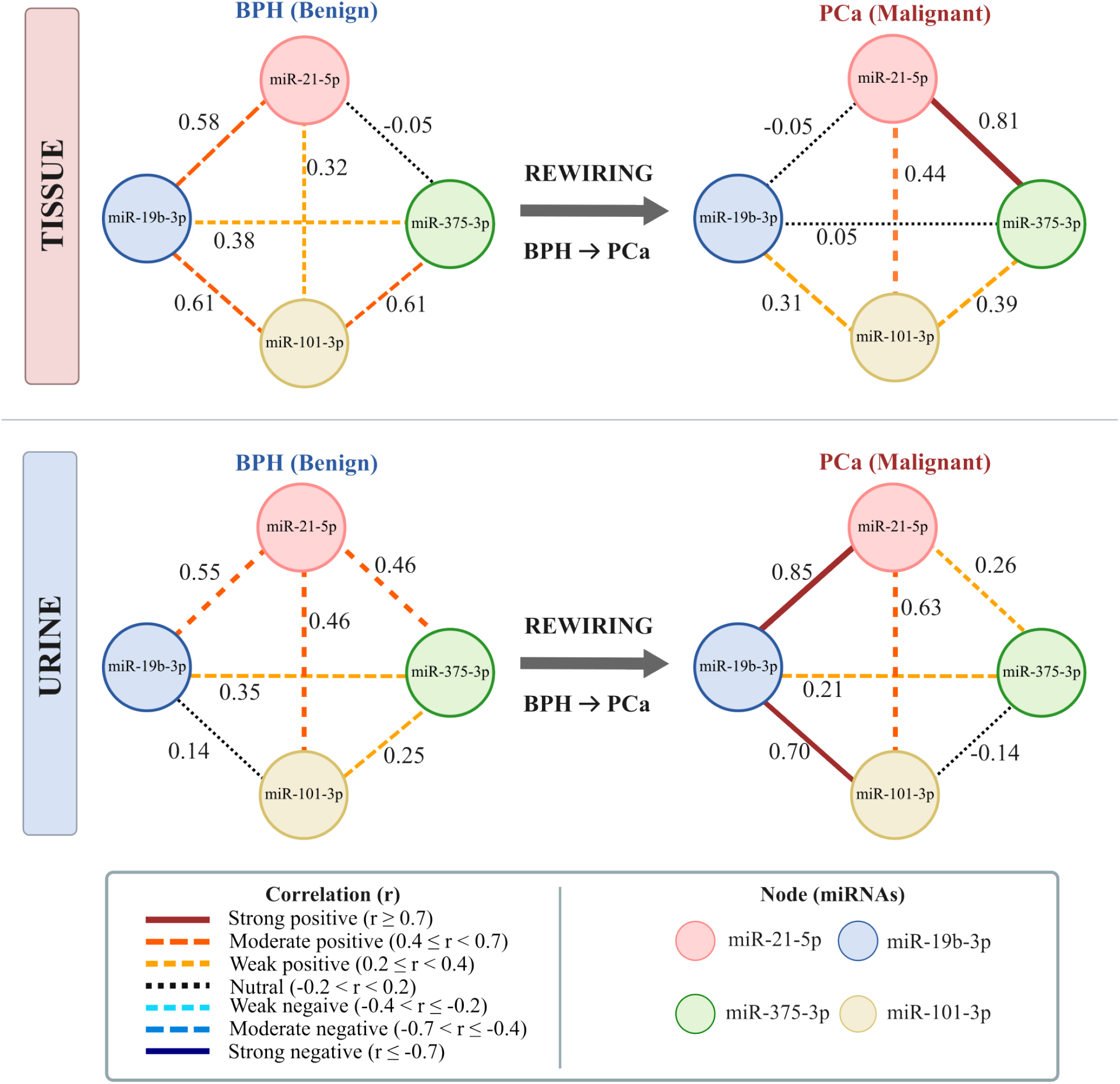
Disease-associated rewiring of miRNA coordination networks in tissue and urine. Network diagrams showing pairwise Spearman correlations among the four candidate miRNAs in BPH and PCa samples. Node colors represent individual miRNAs. Edge color and line style indicate correlation strength and direction.

To express the overall magnitude of coordination change in tissue as a single summary measure, we calculated a NRS. A formal comparison of network architecture revealed substantial miRNA rewiring in both tissue and urine. NRS was greater in tissue (NRS = 2.464) than in urine (NRS = 1.774), indicating that the transition from BPH to PCa is accompanied by more extensive reorganization of studied miRNA coordination within the tumor tissue and urine.

### 4.4 Urine’s Coordination Architecture Is Not a Downstream Reflection of Tumor Tissue Organization

The miR-21-5p–miR-375-3p pair, which underwent the largest coordination change of any pair in tissue (Δ*r* = +0.87), showed a weak negative correlation in urine (Δ*r* ≈ −0.20). The two compartments do not merely differ in magnitude here but they point in opposite directions (Tab. 2). This pair shows strong correlation in PCa tissues as compared to BPH tissue; while the same pair showed negligible correlation in both PCa and BPH urine samples.

The miR-21-5p–miR-19b-3p pair shows no correlation PCa tissue and sight positive correlation BPH tissue; However in urine samples the same pair showed strong positive correlation (*r* = 0.85) in PCa cases and mild positive in BPH cases. Showing the exact opposite pattern as compared to tissue.

The miR-19b-3p–miR-375-3p pair showed the closest alignment of the pairs examined, with a moderate negative shift in tissue (Δ*r* = −0.33) and a weak negative correlation in urine (*r* ≈ −0.14), directionally consistent, though substantially smaller in magnitude in urine.

Most notably, urine’s dominant coordination axis miR-101-3p paired with miR-19b-3p has no corresponding strong relationship in tissue at all; it appears to be an architecture that emerges in urine rather than one that is carried over from the tumor.

To determine whether urinary network alterations reflected those observed in tissue, a Cross-Compartment Concordance Score (CCCS). The resulting score was negative (CCCS = −0.392), indicating that the direction and magnitude of rewiring in urine were largely discordant with those observed in tissue. Thus, although both compartments exhibited disease-associated changes in miRNA coordination, the specific patterns of network reorganization differed substantially and urine do not directly recapitulate the correlation structure of primary tumor tissue (Fig.3).

### 4.5 Urinary miR-101-3p Improves Diagnostic Specificity Beyond PSA

Previous sections established urine as the most informative compartment on both expression and coordination axes, with miR-101-3p emerging as the single strongest marker (Fig. 2). We next asked whether this marker provides diagnostic value when evaluated against the standard clinical comparator PSA in the full cohort (45 urine samples, 28 PCa and 17 BPH) using LOOCV on selected ML-models.

At the conventional PSA cutoff of 4 ng/mL, sensitivity was 100% but specificity was only 23.5%. In practical terms, this cutoff correctly identified every PCa case but flagged the large majority of BPH patients as positive, reflecting the diagnostic gap as described in the Introduction. Only miRNA-101-3p, the LR and EN both get the AUC of 0.71 (95% CI: 0.56–0.85) with 23.5% specificity at 90% sensitivity criteria, similar to the PSA specificity. Which shows no incremental discriminatory value and miRNA-101-3p standalone was clinically unstable(Fig. 4).

**Figure 4:**
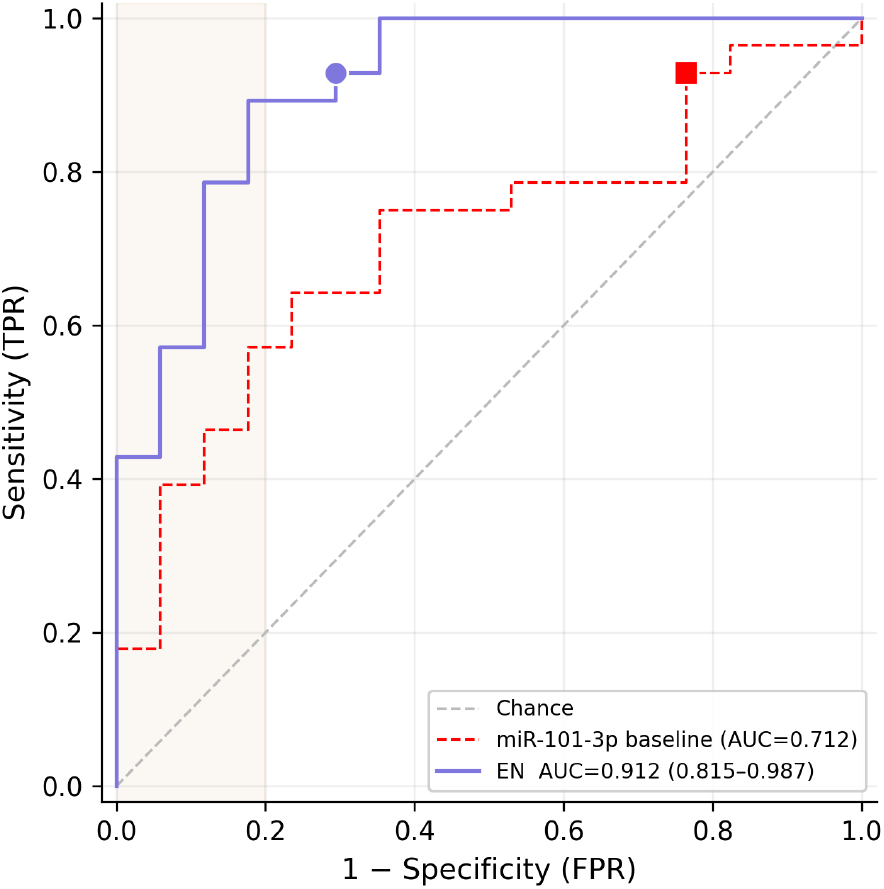
Diagnostic performance for distinguishing prostate cancer from benign prostatic hyperplasia (BPH). The red curve represents the receiver operating characteristic (ROC) curve for urinary exosomal miR-101-3p used as a standalone biomarker. The blue curve represents the ROC curve for a combined model incorporating urinary exosomal miR-101-3p, serum PSA, and age. Area under the curve (AUC) values with corresponding 95% confidence intervals are provided in the legend.

Adding PSA and age improved discrimination beyond mir-101-3p alone. The EN model achieved the highest performance, with an AUC of 0.91 (95% CI: 0.82–0.99) with 70.5% specificity at 92.8% sensitivity. LR and NB showed a similar trend, achieving an AUC of 0.90 (95% CI: 0.82–0.99) and 0.89 (95% CI: 0.76–0.98), with specificity of 70.5% and 76.5% respectively both at 90% sensitivity criteria. These results indicate that miR-101-3p contributes information beyond PSA and improves the separation between prostate cancer and control samples (Suppl. Tab. 2).

Expanding the feature set to include PSA, age, all four urinary miRNAs, and four pair-derived features (i.e. psa, age, urn_21-5p_ct, urn_19b-3p_ct, urn_mir-375-3p_ct, urn_101-3p_ct, pair_diff_19_101, pair_diff_375_101, pair_ratio_19_101, pair_ratio_375_101) did not improve performance beyond the parsimonious model EN AUC of 0.88 (95% CI: 0.77–0.97) (Suppl. Fig. 2 (b)). These findings suggest that most of the discriminative signal was already captured by PSA and miR-101-3p, while the additional variables provided limited incremental benefit in this cohort.

## 5 Discussion

This study examined the diagnostic and network-level properties of four candidate miRNAs across four biological compartments, tissue, serum, whole blood, and urinary exosomes in a cohort of patients undergoing prostate biopsy for clinical suspicion of prostate cancer. Three observations emerge from the data with reasonable consistency. First, the magnitude and direction of cancer-associated miRNA expression changes are compartment-dependent and cannot be predicted from one compartment to another. Second, prostate cancer is accompanied by substantial reorganization of inter-miRNA coordination in both tumor tissue and urine, and these reorganization patterns are largely discordant between the two compartments. Third, urinary miR-101-3p, when combined with PSA, improves diagnostic specificity substantially over PSA used alone.

The most consistent theme across the expression analysis is that miRNA biomarker behavior is not a stable and it is a property of the molecule within a specific biological matrix. Several miRNAs showed not only quantitative differences across compartments but directional reversals in effect size between PCa and BPH. miR-19b-3p showed positive effects in tissue and serum but negative effects in urine and blood. miR-375-3p showed a negative effect in tissue and a positive effect in serum. This is a practically important finding for biomarker development because it means that the classification framework established in one compartment including the direction and threshold cutoff does not translate directly to another.

Compartment-specific miRNA profiles are not unexpected. Circulating miRNAs in serum and plasma exist in multiple physical forms, including Argonaute-2 protein complexes, high-density lipoprotein particles, and membrane-enclosed vesicles, and their relative representation differs between these fractions [21, 22]. Urinary exosomal miRNAs, by contrast, reflect a specific vesicular cargo derived in part from cells lining the urinary tract and, in the case of post-DRE urine, from prostatic secretions. The cellular sources, packaging mechanisms, and degradation kinetics are different in each matrix, and there is no *a priori* reason to expect that the net direction of change observed in a tumor biopsy will be preserved through these successive biological filters.

What is notable in the present data is that blood showed uniformly small effect sizes across all four miRNAs (|*d*| *<* 0.25). The limited discriminatory performance observed in whole blood may be partly because cellular miRNA from hematopoietic cells dominates the signal and partly because the tumor-to-whole-blood volume ratio dilutes any cancer-derived contribution. Serum and tissue showed marker-dependent and inconsistent diagnostic performance, with no single marker achieving acceptable discrimination across both. Only urine provided consistent discrimination across multiple markers, which is biologically plausible given the proximity of urine samples to the prostatic lumen.

An important caveat must be registered here. The RNA isolation protocols were not uniform across compartments: TRIzol-chloroform extraction was used for blood, serum, and tissue, while a commercial exosome isolation kit was used for urine. Differential recovery efficiency across these methods could contribute to apparent compartment differences in expression, independent of genuine biological variation. The present study cannot fully disentangle technical from biological sources of compartment-specificity, and this limitation should be resolved in future work through matched-extraction comparisons or spike-in controls for isolation efficiency.

Beyond expression levels, this study examined whether pairwise correlations among the four candidate miRNAs differed between PCa and BPH. The most pronounced shift was the miR-21-5p/miR-375-3p pair in tumor tissue, where a near-zero correlation in BPH (*r* = −0.05) became strongly positive in PCa (*r* = +0.81; Δ*r* = +0.87). A second substantial shift was observed for miR-19b-3p/miR-101-3p in urine (Δ*r* = −0.64). The aggregate magnitude of these changes, quantified as the Network Rewiring Score, was larger in tissue (NRS = 2.46) than in urine (NRS = 1.77), consistent with the expectation that primary tumor tissue is the source of the most direct molecular dysregulation.

Differential correlation analysis, the comparison of co-expression relationships between conditions, has been proposed as a complement to differential expression analysis for identifying disease-associated molecular changes that would be missed by examining genes in isolation [23]. The concept is particularly relevant in cancer biology, where coordinated regulatory disruption can occur even when individual marker levels change modestly. The strong co-variation of miR-21 and miR-375 specifically in PCa tissue is biologically plausible. miR-21-5p is among the most consistently upregulated miRNAs across human solid tumors, targeting multiple tumor suppressor pathways including PTEN and PDCD4 [24, 25]. miR-375 has well-characterized roles in the prostate, where it targets PTPN4, that functions upstream of STAT3 enhancing the expression of EMT markers and AR but suppressed apoptosis markers [26]. Under malignant conditions, the functional convergence of these two miRNAs on overlapping downstream targets could explain the observed co-variation. However, this interpretation requires validation in a larger, independent tissue cohort before mechanistic conclusions can be drawn.

Perhaps the most conceptually interesting observation in this study is the negative CCCS = −0.392 between tissue and urine, indicating that the direction and magnitude of inter-miRNA rewiring in urinary exosomes is largely discordant from that in primary tumor tissue. Specifically, the miR-21-5p/miR-375-3p pair which showed the largest coordination change in tissue showed a weak inverse pattern in urine. Conversely, the dominant coordination axis in urine (miR-101-3p paired with miR-19b-3p) has no counterpart relationship in tissue. The two compartments thus appear to undergo independent patterns of miRNA network reorganization in the context of malignancy.

This finding challenges an implicit assumption that has shaped the liquid biopsy field: that circulating or excreted biomarkers carry an attenuated version of the tumor’s molecular state, and that the primary analytical task is to detect this signal above background noise. The data presented here, if confirmed in larger cohorts, suggest that urinary exosomal miRNA relationships are shaped by processes specific to the urinary compartment rather than by passive transmission of tumor-intrinsic regulatory structure.

Candidate processes include selective sorting of miRNAs into exosomes during vesicle biogenesis, which is influenced by RNA sequence motifs recognized by heterogeneous nuclear ribonucleoproteins [27], differential stability during transit through the lower urinary tract, and contributions from urothelial or stromal cells resident in the urinary tract that may themselves respond to paracrine signals from the tumor microenvironment.

The current clinical standard for prostate cancer early detection, PSA, is characterized by high sensitivity but poor specificity at the conventionally used threshold of 4 ng/mL. In this cohort, the 4 ng/mL PSA cutoff achieved 100% sensitivity but only 23.5% specificity among 45 urine-matched samples, meaning the large majority of BPH patients with elevated PSA would proceed to biopsy under standard criteria. This reflects the well-documented diagnostic gap: approximately 70–75% of men who undergo biopsy due to elevated PSA do not have cancer [28].

The Elastic Net model combining PSA and urinary miR-101-3p achieved an AUC of 0.91 (95% CI: 0.82–0.99) under leave-one-out cross-validation in this cohort. This represents a meaningful improvement in specificity over PSA alone, and the direction of the finding is consistent with prior reports of urinary miR-101 in prostate cancer. miR-101-3p has been shown to target EZH2, a histone methyltransferase that is overexpressed in high-grade prostate cancer and whose suppression correlates with disease progression [29]. The presence of miR-101-3p in urinary exosomes may therefore reflect active vesicle-mediated export of a tumor suppressor miRNA from prostate epithelial cells, though whether this represents a functional or incidental cargo remains uncertain.

Several limitations qualify the strength of this diagnostic result. The cohort comprised 45 urine samples with a 28:17 PCa:BPH class ratio. LOOCV estimates of AUC can be optimistically biased at this sample size, particularly when feature selection and model fitting are performed within the same validation loop. The wide confidence interval (0.82–0.99) reflects this uncertainty directly, and the lower bound of 0.82 is the more conservative and informative estimate. The finding requires prospective validation in an independent cohort before it can be used to make claims about clinical utility. Also, comparison with existing urinary biomarker tests for prostate cancer including PCA3 (PROGENSA), SelectMDx, and the ExoDx Prostate IntelliScore in the same patient population could add more insight on whether miR-101-3p adds information beyond what is already available through validated tools.

Notably, expanding the feature set beyond PSA and miR-101-3p to include all urinary miRNAs and pairwise features did not improve model performance. This suggests that, in this cohort, the incremental information contributed by the additional markers was limited. Whether this reflects genuine redundancy, insufficient sample size to detect marginal contributions, or the absence of multi-marker benefit in this patient population is not determinable from the present data.

Several structural limitations of the study should be considered alongside the findings. The sample sizes are and the study was conducted at a single centre. Results may not generalize to other populations. The use of histopathological diagnosis on 12-core biopsy as the reference standard may introduce sampling error inherent to that procedure, particularly for low-grade, spatially heterogeneous tumors.

Future work should include prospective validation in an independent cohort, matched extraction controls to separate technical from biological compartment effects, and extension of the coordination analysis to a larger miRNA panel to determine whether the observed network rewiring patterns are reproducible and whether the CCCS framework generalizes to other biofluid pairings. The relationship between urinary exosomal miRNA coordination patterns and Gleason grade or pathological staging would also be informative, as it would help clarify whether the network-level changes observed here are specific to malignancy broadly or scale with disease severity.

## 6 Conclusion

This study provides evidence that miRNA expression profiles and inter-miRNA coordination patterns in prostate cancer are specific to the biological compartment in which they are measured, and that urinary exosomes represent the most diagnostically informative non-invasive compartment among those examined. The observation that urinary miRNA coordination architecture does not recapitulate the tissue coordination structure suggests that liquid biopsy signals carry compartment-specific information rather than a simple diluted reflection of the primary tumor. Urinary miR-101-3p improves discrimination between PCa and BPH beyond PSA in this cohort, though the small sample size requires independent validation before clinical implications can be drawn. Together, these findings argue that the design of miRNA-based liquid biopsy assays should account for the matrix-dependent nature of both individual marker behavior and inter-marker relationships.

## Supporting information

revised_supplemental (supplemental pdf, .tex, figures)

## Data Availability

All data produced in the present study are available upon reasonable request to the authors

## Author Contributions

Research conceptualization and design: Jain G; Supervision and Validation: Jain G,Trivedi S;Project Administration:Jain G; Collection and/or assembly of data: Singh S; Data analysis and interpretation:Jain G, Biswas P, Gupta M; Writing-Original Draft: Jain G, Singh S, Biswas P;Resources: Yadav M, Kumar U, Singh Y, Trivedi S and Das P;Writing-Review & Editing: Jain G,Biswas P,Singh S; Funding Acquisition: Jain G

## Ethics approval and consent to participate

This study was approved by the Ethics Committee of Institute of Science, BHU. The written informed consent was obtained from each participant prior to sampling.

## Data Availability Statement

The datasets generated and/or analyzed in this study are not publicly available due to patient privacy and ethical restrictions but are available from the corresponding author upon reasonable request and subject to approval by the relevant institutional ethics committee.

## Acknowledgement

We would like to express our gratitude to the Department of Urology, IMS, BHU, Varanasi, for their assistance with sample collection, clinical evaluation, and patient recruitment. We are also grateful to the Department of Computer Science, Center for Genetic Disorders and Molecular Human Genetics, Institute of Science, BHU, for providing the lab instruments, technical assistance, and computational expertise required for this research. We would like to express our gratitude to Dr. Madan Gopal Bhardwaj for all of his help to provide clinical resources and patients sampling. Above all, we would like to express our sincere gratitude to the patients and their families for their cooperation, involvement, and trust without which this study would not have been possible.

## Abbreviations

AUC: Area Under Curve
BPH: Benign prostatic hyperplasia
CI: Confidence Interval
CCCS: Cross-Compartment Concordance Score
DRE: Digital rectal examination
LOOCV: Leave-one-out cross-validation
miRNAs: MicroRNAs
NRS: Network Rewiring Score
PCa: Prostate cancer
PSA: Prostate Specific Antigen
qRT-PCR: Quantitative Reverse Transcription Polymerase Chain Reaction
ROC: receiver operating characteristic curve
LR: Logistic Regression
EN: Elastic Net Logistic Regression
NB: Naive Bayes

---

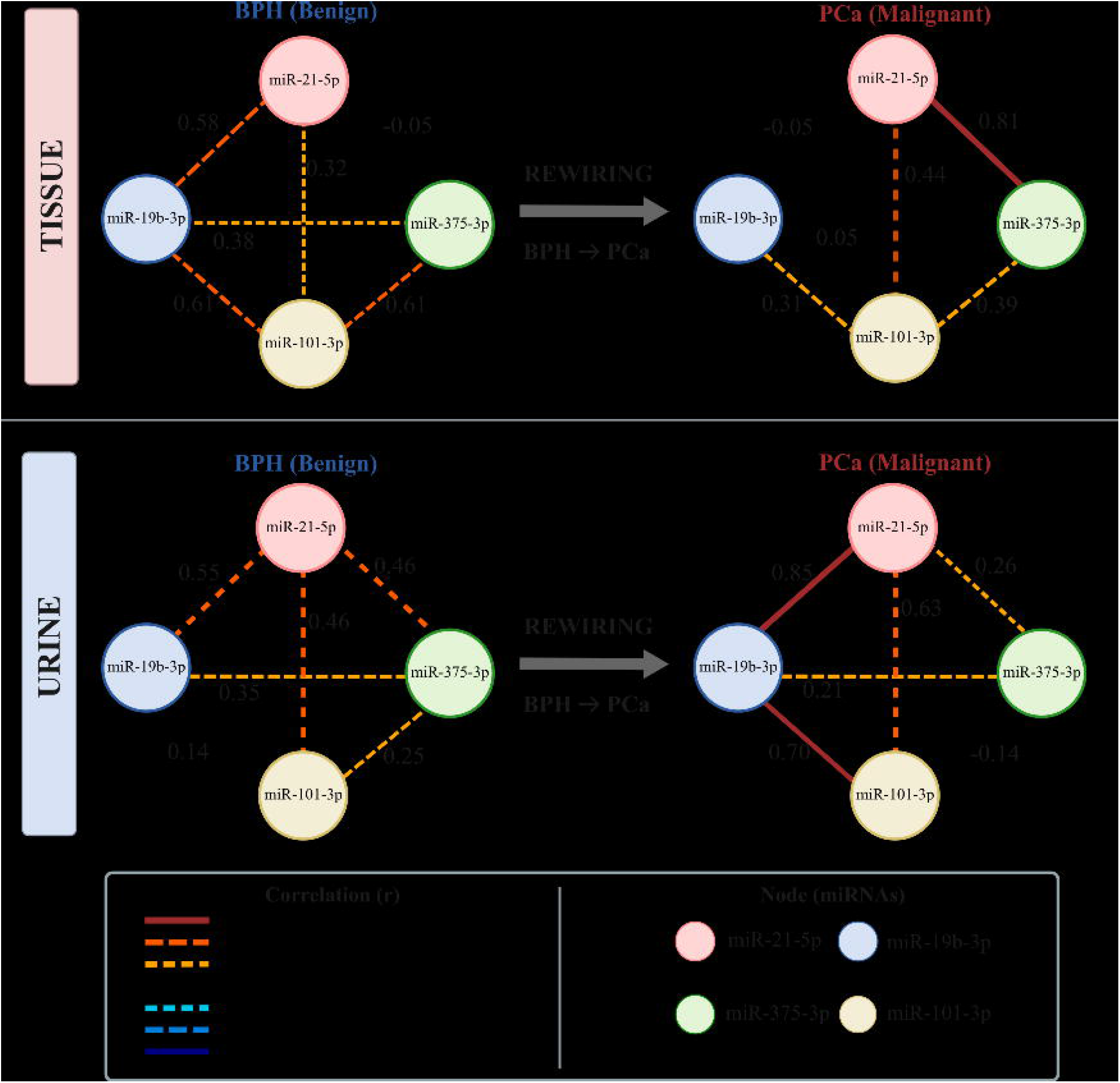

