## Supplementary material for "What Urine Measures Is Not What Tissue Encodes: Compartment-Specific miRNA Coordination in Prostate Cancer": revised_supplemental (supplemental pdf, .tex, figures): supplementary.tex

\documentclass[10pt, a4pages]{article}
\usepackage{graphicx}
\usepackage{amsmath}
\usepackage{subcaption}
\usepackage{geometry}
\geometry{
     top =  1in,
     bottom =  1in,
     right = 1in,
     left = 1in,
}

\renewcommand{\figurename}{Supplementary Figure}
\renewcommand{\tablename}{Supplementary Table}

\title{Supplementary for \textbf{What Urine Measures Is Not What Tissue Encodes: Compartment-Specific miRNA Coordination in Prostate Cancer}}
\date{}
\begin{document}
\maketitle

\section{Methods}
\begin{table}[!ht]
    \centering
    \begin{tabular}{|l|l|l|}
    \hline
        \textbf{Oligo Name} & \textbf{5'-----Sequence-----3'} & \textbf{Length} \\ \hline
        miR-21-5p(SL) & GTCGTATCCAGTGCAGGGTCCGAGGTATTCGCACTGGATACGACTCAACA & 50bp \\ \hline
        miR-21-5p(FP) & ACCACCGTAGCTTATCAGACT & 21bp \\ \hline
        miR-19B-3P(SL) & GTCGTATCCAGTGCAGGGTCCGAGGTATTCGCACTGGATACGACTCAGTT & 50bp \\ \hline
        miR-19B-3P(FP) & AACCGGTGTGCAAATCCATG & 20bp \\ \hline
        miR-101-3P(SL) & GTCGTATCCAGTGCAGGGTCCGAGGTATTCGCACTGGATACGACTTCAGT & 50bp \\ \hline
        miR-101-3P(FP) & AAGCGCCTTACAGTACTGTGA & 20bp \\ \hline
        miR-375-3P(SL) & GTCGTATCCAGTGCAGGGTCCGAGGTATTCGCACTGGATACGACTCACGC & 50bp \\ \hline
        miR-375-3P(FP) & AACCGGTTTGTTCGTTCGG & 19bp \\ \hline
        UniversalRevQ & CCAGTGCAGGGTCCGAGGTA & 20bp \\ \hline
        RNU6 FP & CTCGCTTCGGCAGCACA & 17bp \\ \hline
        RNU6 RP & AACGCTTCACGAATTTGCGT & 20bp \\ \hline
    \end{tabular}
    \caption{Primer Sequence list used for amplification of miRNA and endogenous control.}
    \label{sup-tab-1}
\end{table}
\subsection{Statistical analysis}
Cohen’s d provides a standardized measure of the difference between two group means and is defined as:

\begin{equation}
    d = \frac{\mu_{PCa} - \mu_{BPH}}{s_p}
\end{equation}

where $\mu_{PCa}$ and $\mu_{BPH}$ denote the mean expression values for PCa and BPH groups, respectively, and $s_p$ is the pooled standard deviation given by:

\begin{equation}
    s_p = \sqrt{\frac{(n_{PCa}-1)s^2_{PCa} + (n_{BPH}-1)s^2_{BPH}}{n_{PCa} + n_{BPH} - 2}}
\end{equation}

where $s^2_{PCa}$ and $s^2_{BPH}$ represent the variances of the two groups, and $n_{PCa}$ and $n_{BPH}$ denote their respective sample sizes.\\
Spearman's rank correlation coefficient ($\rho$ or $r$) was calculated as

\begin{equation}
r = 1 - \frac{6\sum_{i=1}^{n} d_i^2}{n(n^2 - 1)}
\end{equation}

where $d_i$ is the difference between the ranks of paired observations and $n$ is the total number of observations.
Differential correlation was quantified as

\begin{equation}
\Delta r = r_{\mathrm{PCa}} - r_{\mathrm{BPH}},
\end{equation}

where $r_{\mathrm{PCa}}$ and $r_{\mathrm{BPH}}$ denote the Spearman
correlation coefficients in prostate cancer and benign prostatic
hyperplasia samples, respectively. To summarize global network
reorganization, a Network Rewiring Score (NRS) was computed as

\begin{equation}
\mathrm{NRS}
=
\frac{1}{E}
\sum_{i=1}^{E}
\left|
r_{i,\mathrm{PCa}}
-
r_{i,\mathrm{BPH}}
\right|,
\end{equation}

where $E$ is the total number of miRNA--miRNA interactions considered.
Higher NRS values indicate greater rewiring of the correlation network
between conditions.

This metric uses the set of pairwise correlation differences as input and produces a single positive value that reflects the overall magnitude of network reorganization within a compartment, with larger values indicating greater disruption of miRNA coordination.\\
The Cross-Compartment Concordance Score (CCCS) was computed using cosine similarity between the vectorized edge-weight profiles of two biological compartments:

\begin{equation}
\mathrm{CCCS}
=
\frac{\mathbf{x}\cdot\mathbf{y}}
{\|\mathbf{x}\|\|\mathbf{y}\|}
=
\frac{\sum_{i=1}^{n} x_i y_i}
{\sqrt{\sum_{i=1}^{n} x_i^2}
 \sqrt{\sum_{i=1}^{n} y_i^2}}
\end{equation}

where:

\begin{itemize}
    \item $\mathbf{x}=(x_1,x_2,\ldots,x_n)$ and $\mathbf{y}=(y_1,y_2,\ldots,y_n)$ are the vectors of edge weights (e.g., miRNA--miRNA correlations) from the two compartments being compared.
    \item $\mathbf{x}\cdot\mathbf{y}$ denotes the dot product between the two vectors.
    \item $\|\mathbf{x}\|$ and $\|\mathbf{y}\|$ denote the Euclidean norms of the vectors.
    \item $n$ is the number of shared miRNA pairs (network edges) considered in the comparison.
\end{itemize}

When signed correlation values are retained, the score satisfies

\begin{equation}
-1 \leq \mathrm{CCCS} \leq 1.
\end{equation}

The interpretation of CCCS is as follows:

\begin{itemize}
    \item $\mathrm{CCCS} \approx 1$: highly concordant network organization across compartments.
    \item $\mathrm{CCCS} \approx 0$: little or no similarity in correlation structure.
    \item $\mathrm{CCCS} \approx -1$: opposite network organization, where positive relationships in one compartment tend to be negative in the other.
\end{itemize}
A data-driven quantile-based thresholding strategy was applied to select informative gene pairs. Based on the distribution of $\left|\Delta r\right|$, gene pairs were categorized as: strong pairs ($\left|\Delta r\right| \geq Q75$), very strong pairs ($\left|\Delta r\right| \geq Q90$). 

\begin{figure}
    \centering
    \begin{subfigure}[b]{0.48\textwidth}
        \centering
        \includegraphics[width=0.5\linewidth]{supplementary/sup_fig2/sup-tis.png}
        \caption{Tissue}
    \end{subfigure}
    \hfill
    \begin{subfigure}[b]{0.48\textwidth}
        \centering
        \includegraphics[width=0.5\linewidth]{supplementary/sup_fig2/sup-tis.png}
        \caption{Serum}
    \end{subfigure}
    \hfill
    \begin{subfigure}[b]{0.48\textwidth}
        \centering
        \includegraphics[width=0.5\linewidth]{supplementary/sup_fig2/sup-bld.png}
        \caption{Blood}
    \end{subfigure}
    \caption{\textit{Diagnostic performance of miRNAs for distinguishing prostate cancer from benign prostatic hyperplasia. Receiver operating characteristic (ROC) curves showing the classification performance of individual (a)Tissue, (b)Serum and (c) Blood miRNAs. Area under the curve (AUC) values and corresponding 95\% confidence intervals are shown in the legend.}}
    \label{fig:placeholder}
\end{figure}

\newpage
\section{Results}


\begin{table}[!ht]
    \centering
    \begin{tabular}{|l|l|l|l|l|l|l|l|}
    \hline
        \textbf{Model} & \textbf{Acc} & \textbf{Sen} & \textbf{Spe.} & \textbf{PPV} & \textbf{NPV} & \textbf{AUC} & \textbf{AUC 95\% CI} \\ \hline
        PSA $\geq$ 4 ng/mL (rule) & 0.7111 & 1.0 & 0.2353 & 0.6829 & 1.0 & 0.8939 & 0.793–0.976 \\ \hline
        ct 101 only (LR) & 0.6667 & 0.9286 & 0.2353 & 0.6667 & 0.6667 & 0.7122 & 0.558–0.850 \\ \hline
        ct 101 only (EN) & 0.6667 & 0.9286 & 0.2353 & 0.6667 & 0.6667 & 0.7122 & 0.558–0.850 \\ \hline
        ct 101 only (NB) & 0.6222 & 1.0 & 0.0 & 0.6222 & 0.0 & 0.6597 & 0.496–0.816 \\ \hline
        PSA + age + ct miR-101 (LR) & 0.8444 & 0.9286 & 0.7059 & 0.8387 & 0.8571 & 0.9097 & 0.817–0.986 \\ \hline
        PSA + age + ct miR-101 (EN) & 0.8444 & 0.9286 & 0.7059 & 0.8387 & 0.8571 & 0.9118 & 0.815–0.987 \\ \hline
        PSA + age + ct miR-101 (NB) & 0.8667 & 0.9286 & 0.7647 & 0.8667 & 0.8667 & 0.8866 & 0.765–0.981 \\ \hline
        all (LR) & 0.8222 & 0.9286 & 0.6471 & 0.8125 & 0.8462 & 0.8824 & 0.772–0.966 \\ \hline
        all (EN) & 0.8444 & 0.9286 & 0.7059 & 0.8387 & 0.8571 & 0.8845 & 0.772–0.970 \\ \hline
        all (NB) & 0.8 & 0.9643 & 0.5294 & 0.7714 & 0.9 & 0.8697 & 0.755–0.956 \\ \hline
    \end{tabular}
    \caption{ML summary table}
\end{table}
\begin{figure}
    \centering
    \begin{subfigure}[b]{0.48\textwidth}
        \centering
        \includegraphics[width=0.5\linewidth]{supplementary/sup_fig4/(a) miR-101-3p only_auc.png}
        \caption{PSA clinical cut-off only}
    \end{subfigure}
    \hfill
    \begin{subfigure}[b]{0.48\textwidth}
        \centering
        \includegraphics[width=0.5\linewidth]{supplementary/sup_fig4/(c) PSA + 4 miRNA ct + 4 pair (exploratory)_auc.png}
        \caption{PSA, age, 4 miRNAs and 4 difference and ratio features}
    \end{subfigure}
    \caption{\textit{Diagnostic performance of miRNAs for distinguishing prostate cancer from benign prostatic hyperplasia. PSA, age, 4 miRNAs and 4 difference and ratio features. Area under the curve (AUC) values and corresponding 95\% confidence intervals are shown in the legend.}}
    \label{fig:placeholder}
\end{figure}
% \section{Discussion}
\end{document}
