## Supplementary figures and images for "What Urine Measures Is Not What Tissue Encodes: Compartment-Specific miRNA Coordination in Prostate Cancer"

### (a) miR-101-3p only_auc.png

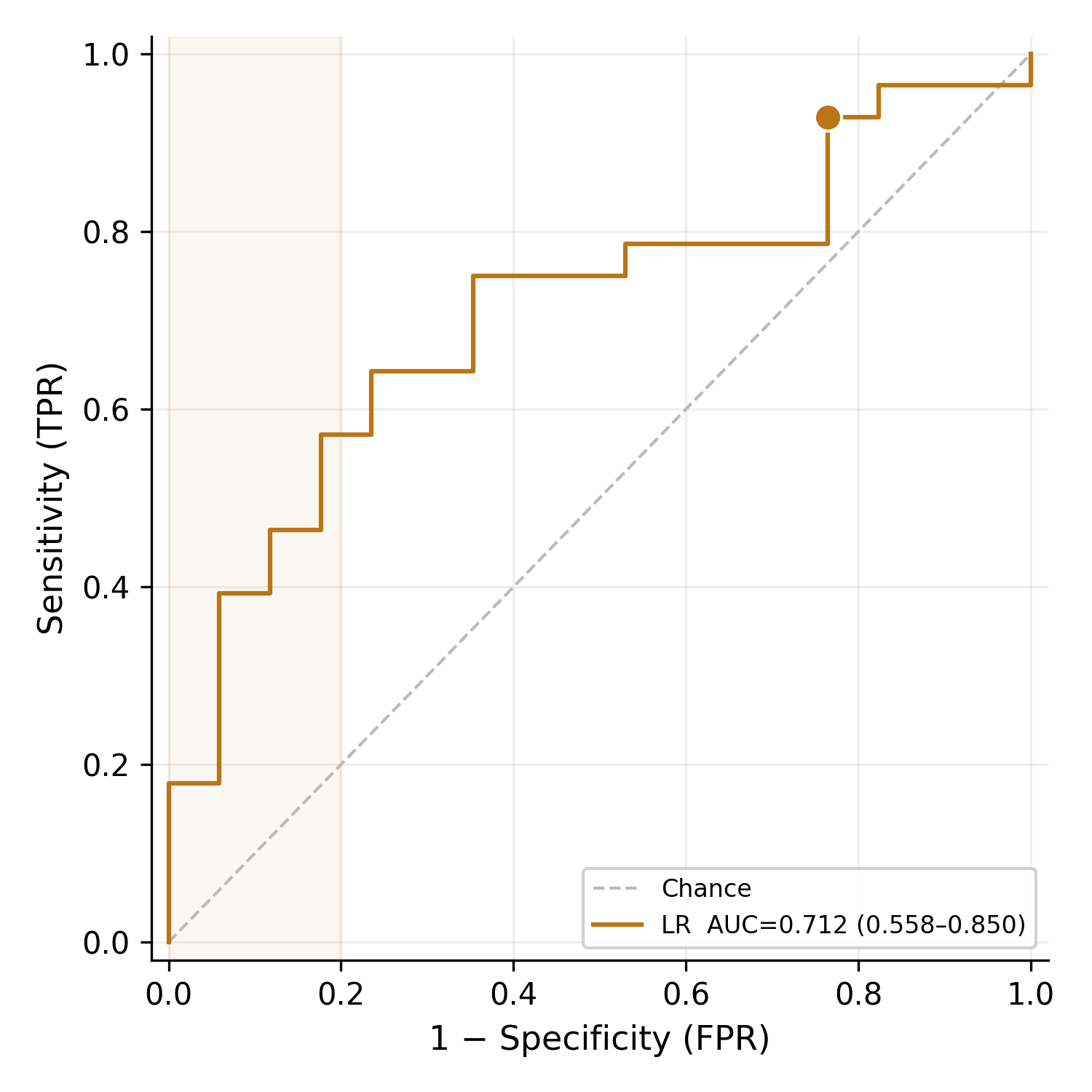

### (c) PSA + 4 miRNA ct + 4 pair (exploratory)_auc.png

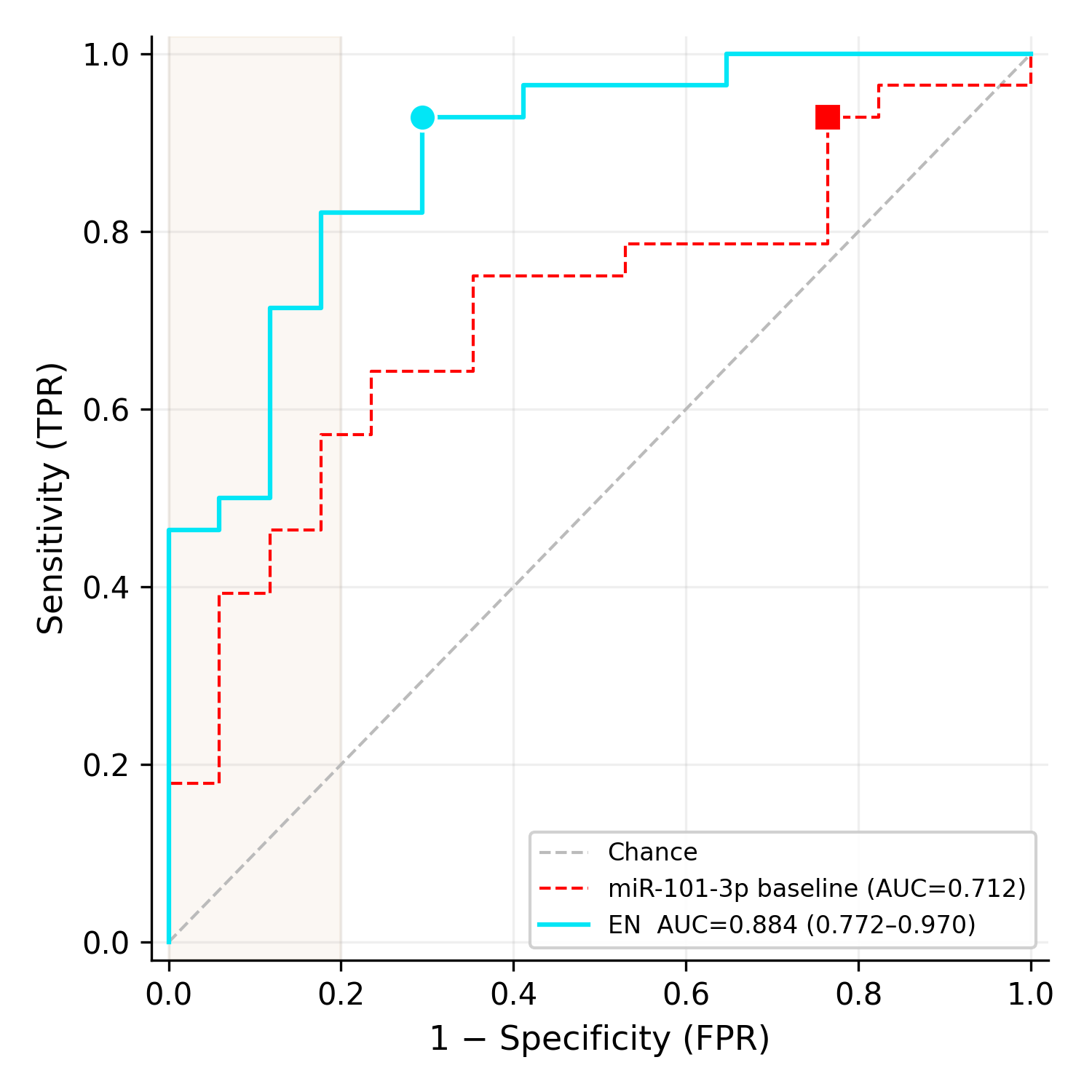

### sup-bld.png

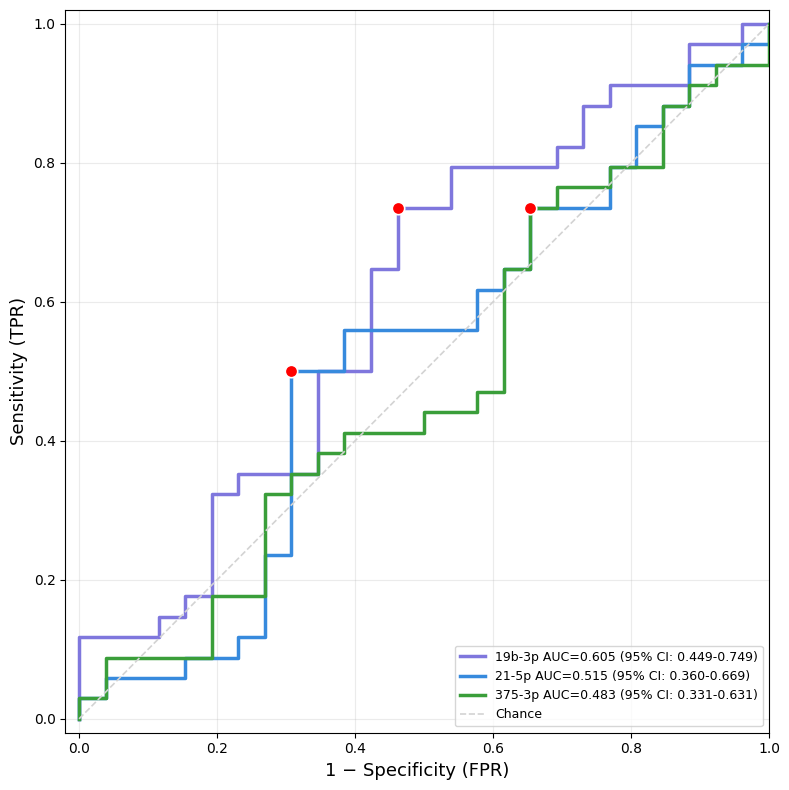

### sup-srm.png

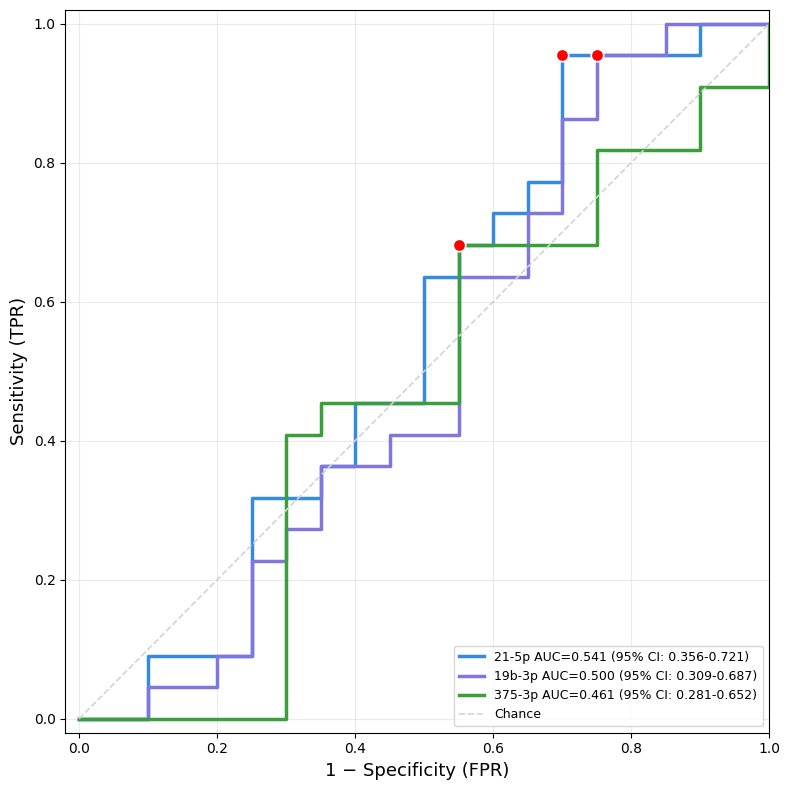

### sup-tis.png

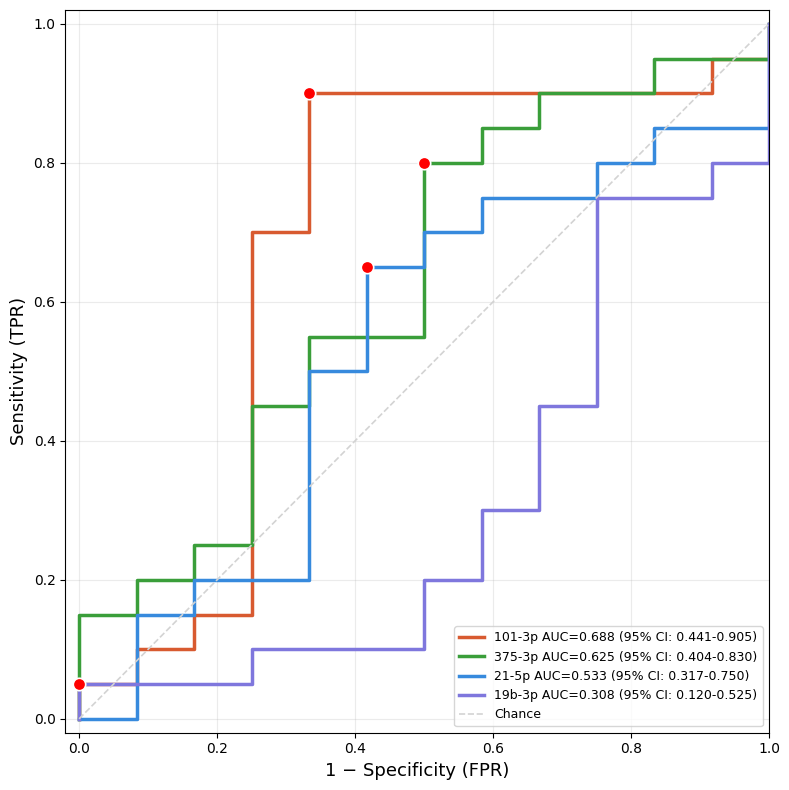
